# Diagnostic performance of a novel digital immunoassay (RapidTesta SARS-CoV-2): a prospective observational study with 1,127 nasopharyngeal samples

**DOI:** 10.1101/2021.07.26.21261162

**Authors:** Hiromichi Suzuki, Yusaku Akashi, Atsuo Ueda, Yoshihiko Kiyasu, Yuto Takeuchi, Yuta Maehara, Yasushi Ochiai, Shinya Okuyama, Shigeyuki Notake, Koji Nakamura, Hiroichi Ishikawa

## Abstract

**Introduction:** Digital immunoassays are generally regarded as superior tests for the detection of infectious disease pathogens, but there have been insufficient data concerning SARS-CoV-2 immunoassays.

**Methods:** We prospectively evaluated a novel digital immunoassay (RapidTesta SARS-CoV-2). Two nasopharyngeal samples were simultaneously collected for antigen tests and RT-PCR. Real-time RT-PCR for SARS-CoV-2, using a method developed by the National Institute of Infectious Diseases, Japan, served as the reference RT-PCR method.

**Results:** During the study period, 1,127 nasopharyngeal samples (symptomatic patients: 802, asymptomatic patients: 325) were evaluated. For digital immunoassay antigen tests, the sensitivity was 78.3% (95% CI: 67.3%–87.1%) and the specificity was 97.6% (95% CI: 96.5%–98.5%). When technicians visually analyzed the antigen test results, the sensitivity was 71.6% (95% CI: 59.9%–81.5%) and the specificity was 99.2% (95% CI: 98.5%–99.7%). Among symptomatic patients, the sensitivity was 89.4% (95% CI; 76.9%–96.5%) with digital immunoassay antigen tests, and 85.1% (95% CI; 71.7%–93.8%) with visually analyzed the antigen test, respectively.

**Conclusions:** The findings indicated that RapidTesta SARS-CoV-2 analysis with the DIA device had sufficient analytical performance for the detection of SARS-CoV-2 in nasopharyngeal samples. When positive DIA results are recorded without a visually recognizable red line at the positive line location on the test cassette, additional RT-PCR evaluation should be performed.

## Introduction

COVID-19 has been the primary global health concern for 2 consecutive years (2020 and 2021) and its diagnosis has been mainly performed by respiratory sampling [1]. RT-PCR has been regarded as the “gold standard” method [1] for the detection of SARS-CoV-2, but a long turnaround time is needed for evaluation of the results.

Immunochromatographic antigen testing is a widely used laboratory technique for the detection of SARS-CoV-2 owing to its ease of handling and rapid acquisition of results [2, 3]; moreover, a strong correlation between antigen test results and SARS-CoV-2 infectivity has been reported [4]. In recent years, improved sensitivities for pathogen detection using digital immunoassay devices have been proven, especially in the field of influenza [5], and applications to COVID-19 diagnosis have been expected.

The RapidTesta SARS-CoV-2 is a novel immunochromatographic assay that can detect SARS-CoV-2 in 10 minutes; it was newly approved in Japan in June 2021. The RapidTesta SARS-CoV-2 can be analyzed using a portal digital immunoassay (DIA) device, with presumably greater sensitivity compared with human-eye judgment.

Herein, we performed a prospective comparison with a DIA device and RT-PCR using a set of two simultaneously collected nasopharyngeal samples from multiple patients.

## Patients and Methods

The current study was prospectively performed between April 20, 2021, and May 31, 2021, at a “drive-through” PCR center in Tsukuba Medical Center Hospital (TMCH), Tsukuba, Japan. During the study period, patients were mainly referred from 63 clinic and a local public health center. Informed consent was obtained from all participating patients and the study was performed with the approval of the TMCH ethical committee (approval number: 2021-021).

### Study process and evaluation of antigen test results with and without the use of a digital immunoassay device

In this study, two nasopharyngeal samples were simultaneously collected with FLOQ Swabs (Copan Italia S.p.A., Brescia, Italy) for RT-PCR and flocked type swabs (contained in the RapidTesta SARS-CoV-2 kit) for antigen tests, as previously described [6]. For antigen tests, each swab was diluted into extraction buffer; three drops of the extracted sample were then added to a test cassette through filtration (Figure 1). The examination time was 10 minutes; the analysis was performed both visually and automatically with a DIA device (Figure 2).

**Figure 1.**
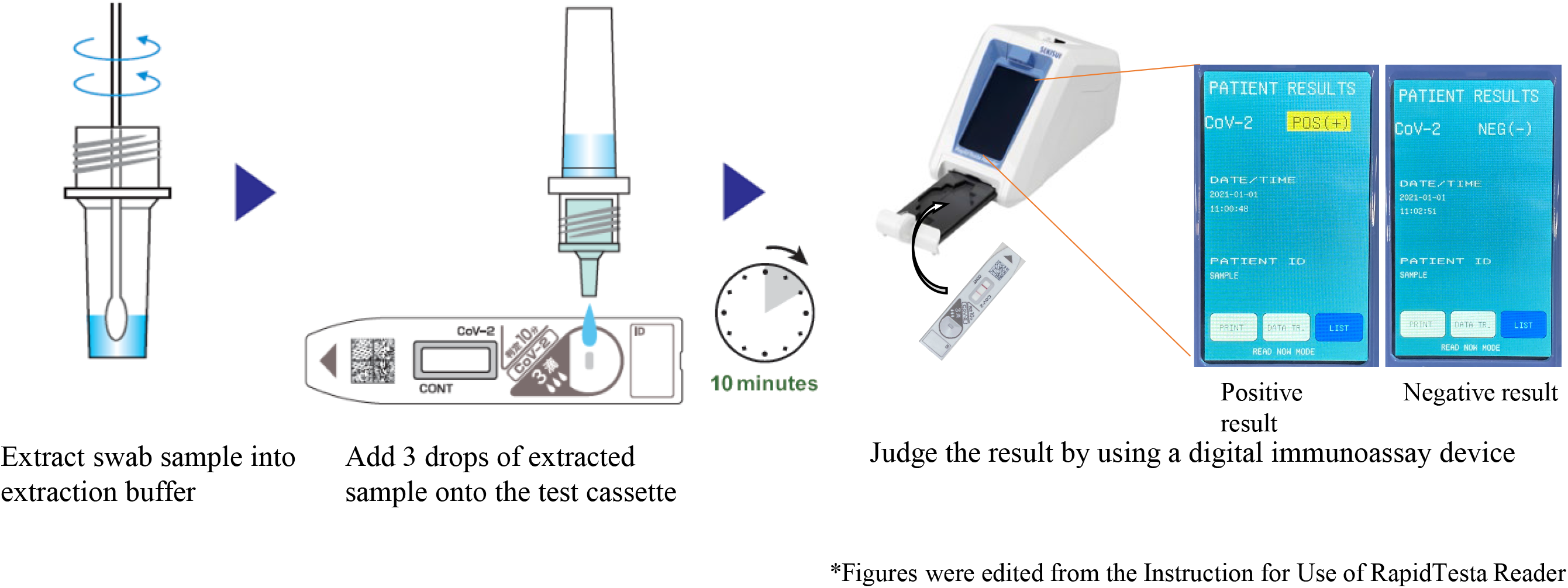
Test flow diagram of the RapidTesta SARS-CoV-2 without the use of a digital immunoassay device

**Figure 2.**
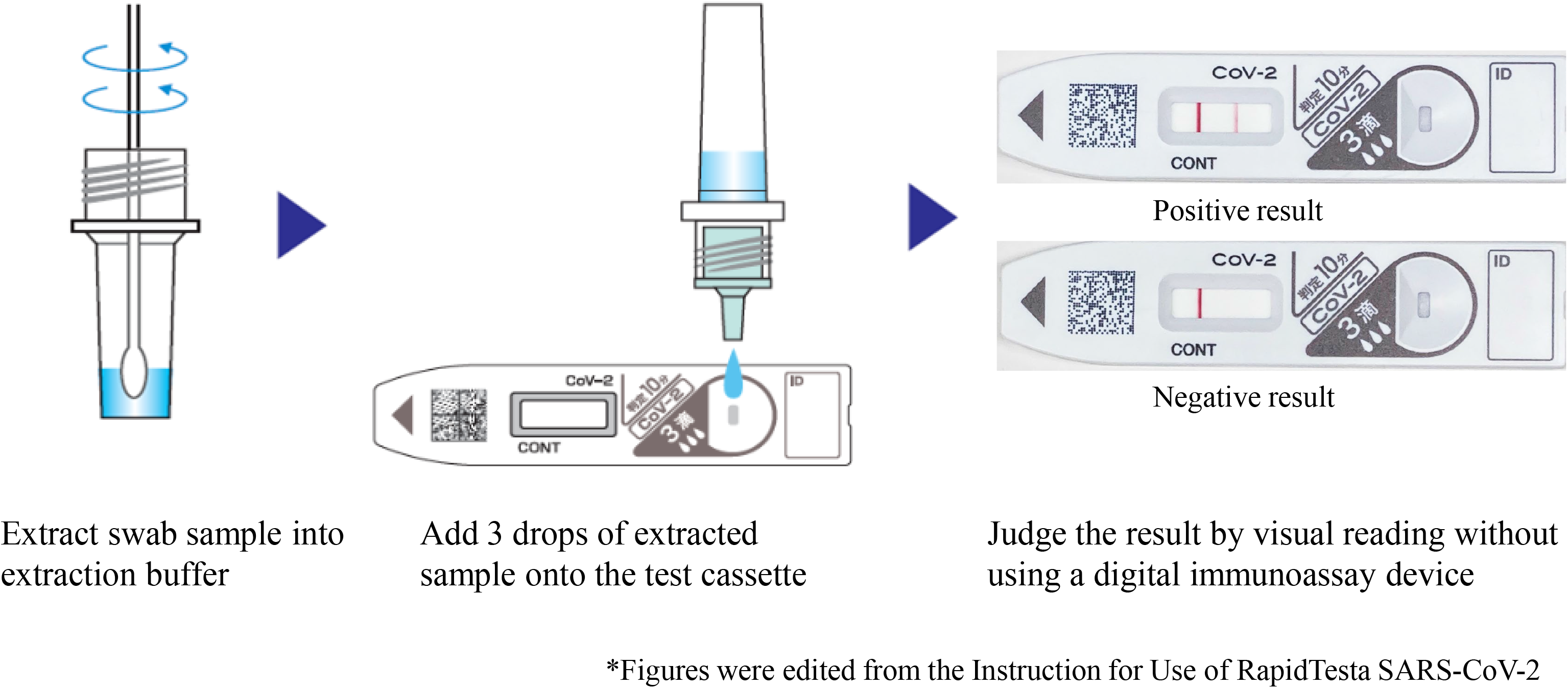
Test flow diagram of the RapidTesta SARS-CoV-2 with the use of a digital immunoassay device

For RT-PCR, each swab was diluted in 3 mL of Universal Transport Medium (BD), then transferred to the microbiology department of the TMCH for RNA extraction and in-house RT-PCR. Extraction was performed with magLEAD 6gC (Precision System Science Co., Ltd., Chiba, Japan) with a 200µL aliquot of each nasopharyngeal sample. In total, 100 µL of purified sample were eluted and subjected to in-house RT-PCR analysis [7] and a reference real-time RT-PCR analysis for SARS-CoV-2, which used a method developed by the National Institute of Infectious Diseases, Japan (NIID method) [8]. The reference RT-PCR analysis was performed at Tsukuba Research Institute of Sekisui Medical Co., Ltd.; eluted samples were transported weekly for evaluation. Before evaluation, all samples were preserved at –80°C.

### Reference RT-PCR using a method developed by the National Institute of Infectious Diseases, Japan

The NIID method has been regarded as the “gold standard” RT-PCR method for SARS-CoV-2 detection and was used as the reference RT-PCR method in this study. The NIID method was performed with duplicate N and N2 assay analysis, as previous described [8]. The CFX96 Touch Real-Time PCR Detection System (Bio-Rad Laboratories, Hercules, CA, USA) and Reliance One-Step Multiplex Supermix (Bio-Rad Laboratories) were used for analysis. In the event of discordance between the reference RT-PCR and in-house RT-PCR results, the GeneXpert® system and an Xpert® Xpress SARS-CoV-2 assay (Cepheid Inc., Sunnyvale, CA, USA) [9] were used for additional evaluation.

### Statistical analysis

The sensitivity, specificity, positive predictive value, and negative predictive value of antigen test results were calculated using the Clopper and Pearson method, with 95% confidence intervals (CIs). All calculations were conducted using R software, version 4.1.0 (www.r-project.org).

## Results

During the study period, 1,127 nasopharyngeal samples were collected for evaluation. In total, 802 samples were collected from symptomatic patients and 325 samples were collected from asymptomatic patients.

Of the 1,127 samples, 74 (6.6%) were positive according to RT-PCR with the NIID method. There were no instances of discordance between RT-PCR with the NIID method and in-house RT-PCR; thus, no additional evaluations with the GeneXpert® system were performed.

The results of the RapidTesta SARS-CoV-2 without the use of a DIA device are shown in Table 1-a. The sensitivity was 71.6% (95% CI: 59.9%–81.5%), and the specificity was 99.2% (95% CI: 98.5%–99.7%). The positive predictive value was 86.9% (95% CI: 75.8%–94.2%), and the negative predictive value was 98.0% (95% CI: 97.0%–98.8%). The results of the RapidTesta SARS-CoV-2 with the use of a DIA device are shown in Table 1-b. The sensitivity was 78.4% (95% CI: 67.3%–87.1%), and the specificity was 97.6% (95% CI: 96.5%–98.5%). The positive predictive value was 69.9% (95% CI: 58.8%–79.5%), and the negative predictive value was 98.5% (95% CI: 97.5%–99.1%).

**Table 1-a.** Sensitivity and specificity of the RapidTesta SARS-CoV-2 without the use of a digital immunoassay device among all patients in the study.

|  |  | real-time RT-PCR |  |
| --- | --- | --- | --- |
|  |  | Positive | Negative |
| Antigen test | Positive | 53 | 8 |
|  | Negative | 21 | 1045 |
| Sensitivity (%) |  | 71.6(59.9-81.5) |  |
| Specificity (%) |  | 99.2(98.5-99.7) |  |

Reverse transcription-polymerase chain reaction findings were used as reference results. RT-PCR, reverse transcription-polymerase chain reaction Data in parentheses indicate 95% confidence intervals.

**Table 1-b.** Sensitivity and specificity of the RapidTesta SARS-CoV-2 with the use of a digital immunoassay device among all patients in the study.

|  |  | real-time RT-PCR |  |
| --- | --- | --- | --- |
|  |  | Positive | Negative |
| Antigen test | Positive | 58 | 25 |
|  | Negative | 16 | 1028 |
| Sensitivity (%) |  | 78.4(67.3-87.1) |  |
| Specificity (%) |  | 97.6(96.5-98.5) |  |

Reverse transcription-polymerase chain reaction findings were used as reference results. RT-PCR, reverse transcription-polymerase chain reaction Data in parentheses indicate 95% confidence intervals.

The results of the RapidTesta SARS-CoV-2 with/without the use of a DIA device are shown in Table 2-a and Table 2-b. For the RapidTesta SARS-CoV-2 without the use of a DIA device, the sensitivity was 85.1% (95% CI: 71.7%–93.8%), and the specificity was 99.3% (95% CI: 98.5%–99.8%). For the RapidTesta SARS-CoV-2 with the use of a DIA device, the sensitivity was 89.4% (95% CI: 76.9%–96.5%), and the specificity was 97.7% (95% CI: 96.4%–98.7%).

**Table 2-a.** Sensitivity and specificity of the RapidTesta SARS-CoV-2 without the use of a digital immunoassay device among symptomatic patients.

|  |  | real-time RT-PCR |  |
| --- | --- | --- | --- |
|  |  | Positive | Negative |
| Antigen test | Positive | 40 | 5 |
|  | Negative | 7 | 750 |
| Sensitivity (%) |  | 85.1(71.7-93.8) |  |
| Specificity (%) |  | 99.3(98.5-99.8) |  |

**Table 2-b.** Sensitivity and specificity of the RapidTesta SARS-CoV-2 with the use of a digital immunoassay device among symptomatic patients.

|  |  | real-time RT-PCR |  |
| --- | --- | --- | --- |
|  |  | Positive | Negative |
| Antigen test | Positive | 42 | 17 |
|  | Negative | 5 | 738 |
| Sensitivity (%) |  | 89.4(76.9-96.5) |  |
| Specificity (%) |  | 97.7(96.4-98.7) |  |

The sensitivities stratified according to Ct value are shown in Table 3. Without the use of a DIA device (Table 3-a), the sensitivity for Ct < 20 was 96.4% (95% CI: 81.7%–99.9%); the sensitivity was 81.3% (95% CI: 63.6%–92.8%) for Ct 20–29. No positive samples (Ct ≥ 30) were detected without the use of a DIA device. When using a DIA device (Table 3-b), the sensitivity for Ct < 20 was 100.0% (95% CI: 87.7%–100%); the sensitivities were 84.4% (95% CI: 67.2%–94.7%) for Ct 20–29 and 21.4% (95% CI: 4.7%–50.8%) for (Ct ≥ 30), respectively.

**Table 3-a.** Sensitivities of the RapidTesta SARS-CoV-2 without the use of a digital immunoassay device according to Ct value.

| Ct value(N2) | Sensitivity (%) | Positive | Negative |
| --- | --- | --- | --- |
| <20 | 96.4(81.7-99.9) | 27 | 1 |
| 20-29 | 81.3(59.0-91.7) | 26 | 6 |
| ≥30 | 0(0-19.5) | 0 | 14 |

**Table 3-b.** Sensitivities of the RapidTesta SARS-CoV-2 with the use of a digital immunoassay device according to Ct value.

| Ct value(N2) | Sensitivity (%) | Positive | Negative |
| --- | --- | --- | --- |
| <20 | 100(87.7-100) | 28 | 0 |
| 20-29 | 84.4(63.1-93.9) | 27 | 5 |
| ≥30 | 21.4(3.8-43.4) | 3 | 11 |

## Discussion

This prospective study revealed that the sensitivity of the RapidTesta SARS-CoV-2 was 78.4% and the specificity was 97.6%. For symptomatic patients, the sensitivities were 85.1% for antigen tests without the use of a DIA device and 89.4% for antigen tests with the use of a DIA device. The sensitivity of DIA tests was superior to the sensitivity of visually analyzed antigen tests, but the rate of false-positive results increased with introduction of a DIA device.

A recent systematic review described overall antigen test sensitivities of 72.0% (95% CI: 63.7%–79.0%) for symptomatic patients and 58.1% (95% CI: 40.2%–74.1%) for asymptomatic patients [10]. For quantitative antigen tests, the overall sensitivity and specificity were 84.8% and 97.9%, respectively [11]. In the current study, the sensitivity for symptomatic patients of RapidTesta SARS-CoV-2 was over 80% and it is considered to meet the acceptable criteria of antigen test recommended by the World Health Organization[12].

To our knowledge, there have been few evaluations of DIA devices for the detection of SARS-CoV-2. Most recently, a prospective study of a DIA device (QuickChaser Auto SARS-CoV-2) with 1,401 nasopharyngeal samples showed that the sensitivity and specificity were 74.7% (95% CI: 64.0%–83.6%) and 99.8% (95% CI: 99.5%–100%), respectively [13]. The sensitivity for positive samples (Ct < 30) was 98.2% (56/57). In our study, the overall sensitivity of the RapidTesta SARS-CoV-2 with a DIA device was comparable with a previous DIA device (QuickChaser Auto SARS-CoV-2). This rapid analysis could be completed within 10 minutes, although its sensitivity for positive samples (Ct < 30) (91.7%; 55/60) was slightly inferior to the previously described DIA device.

False-positive results that indicate the presence of COVID-19 cause substantial harm in clinical practice [14] and the emergence of false-positive results has been repeatedly reported in Japan [15–17]. In the current study, the increased sensitivity when using the DIA device was negatively associated with the decrease of specificity. False-positive results were frequently observed (17/25; 68.0%) via DIA inspection for samples with RT-PCR-negative results in this study. The current study suggested that additional SARS-CoV-2 evaluation should be performed when positive RapidTesta SARS-CoV-2 results with a DIA device are recorded in the absence of a visually recognizable positive line.

There were some limitations to this study. First, study samples were collected in a local district in Japan in the spring of 2021. Second, differences among variant strains were not analyzed. Third, study samples were collected from the nasopharyngeal tract and saliva samples were not analyzed. Fourth, improvements of DIA specificity were not examined and further investigations are necessary.

In conclusion, this study showed that the RapidTesta SARS-CoV-2 with a DIA device had sufficient analytical performance for the detection of SARS-CoV-2 in nasopharyngeal samples. When positive DIA results are recorded without a visually recognizable red line at the positive line location on the test cassette, additional RT-PCR evaluation should be performed.

## Data Availability

The data are not publicly available due to their containing information that could compromise the privacy of research participants.

## Acknowledgements

We thank Mrs. Yoko Ueda, Mrs. Mio Matsumoto, Mr. Masaomi Matsubayashi, Mrs. Mika Yaguchi, Mrs. Yumiko Tanaka, and the staff of the Department of Clinical Laboratory of Tsukuba Medical Center Hospital for their support of this study. We thank all participating medical institutions for providing their patients’ clinical information. Mrs. Yoko Ueda and Mrs. Mio Matsumoto substantially contributed to creating the database for this study.

## Conflict of Interest

Sekisui Medical Co., Ltd. provided fees for research expenses and provided the RapidTesta SARS-CoV-2 without charge. Yuta Maehara, Yasushi Ochiai, and Shinya Okuyama belong to Sekisui Medical Co., Ltd., the developer of the RapidTesta SARS-CoV-2.

## Notes

### Author Declarations

The ethics committee of TMCH approved the present study (approval number: 2021-021).

